# Early gastrointestinal manifestations predict disease progression and mortality in patients with systemic sclerosis

**DOI:** 10.64898/2025.12.01.25341383

**Authors:** Ali Y. Ayla, Ashish P. Balar, Ernesto Calderon Martinez, Bingrui Chen, Claudia Pedroza, Francesco Bonomi, Silvia Bellando-Randone, Maureen D. Mayes, Brian Skaug, Michael Hughes, Shervin Assassi, Zsuzsanna H. McMahan

## Abstract

**Objective:** Gastrointestinal (GI) complications are common in systemic sclerosis (SSc), yet the value of early symptoms as predictors of progression remains poorly defined. We aimed to determine whether baseline GI symptoms in early SSc predict progression to significant GI outcomes.

**Methods:** We studied 450 participants from the GENISOS cohort with early SSc (≤5 years from first non-Raynaud’s symptom). Baseline assessments included demographics and clinical variables, including individual GI symptoms (GERD, dysphagia, bloating, constipation, diarrhea, peptic ulcer). Significant GI involvement was defined as Medsger GI severity score ≥2. Cox models evaluated associations between baseline symptoms and progression to significant GI disease and all-cause mortality. Additional models examined total baseline GI symptom burden and outcomes.

**Results:** Participants were predominantly female (84%) with a mean age of 47.7 ± 13.3 years. Baseline GI symptoms were frequent, most commonly GERD (76%) and dysphagia (41%). 13% of the patients had significant GI involvement, with 4% presenting with significant GI disease at baseline. Baseline GERD (HR 4.15, P=0.019) and diarrhea (HR 2.25, P=0.021) were strong predictors of GI progression. When symptom burden was analyzed, each additional baseline GI symptom increased the hazard of significant GI progression by 53% (P=0.003). On the other hand, peptic ulcer disease (HR 2.21, P=0.007) and diarrhea (HR 1.48, P=0.045) predicted higher mortality, with each additional symptom raising mortality risk by 29% (P<0.001).

**Conclusions:** Early GI symptoms and overall symptom burden predict GI progression and mortality in SSc. Symptom-based profiling may offer a practical strategy for clinical risk stratification and trial enrichment.

**Key messages:** *Key message 1:* Baseline GERD and diarrhea strongly predict progression to significant gastrointestinal involvement in systemic sclerosis.

*Key message 2:* Greater baseline gastrointestinal symptom burden increases risk of severe GI disease progression and all-cause mortality.

*Key message 3:* Simple symptom profiles may guide early risk stratification and patient selection for GI-focused clinical trials.

## Introduction

Systemic sclerosis (SSc) is a multifaceted systemic autoimmune rheumatic disease (SARD) marked by vasculopathy, immune dysregulation, and progressive fibrosis (1). The capacity of SSc to affect nearly every organ system contributes to substantial disease-related morbidity and elevated mortality among affected individuals (2). While significant advances have been made in characterizing and modeling the progression of skin and pulmonary involvement (3-5), gastrointestinal (GI) manifestations, despite being nearly universal in both limited and diffuse SSc, remain comparatively understudied (6). This knowledge gap is particularly concerning, given the profound impact of GI disease on quality of life and nutritional status, as well as the lack of reliable tools to predict its onset, severity, and progression.

SSc can involve any segment of the GI tract—from the oropharynx to the anorectum—with marked variability in both temporal/anatomical presentation and severity. This clinical heterogeneity poses significant challenges in predicting specific GI complications and long-term outcomes. Recent studies suggest that extra-intestinal clinical features may serve as valuable indicators for distinguishing GI phenotypes and anticipating disease trajectories in SSc (7-13). Moreover, several newly identified autoantibodies have been associated with GI involvement, offering promising avenues for stratifying patients by type and severity of GI disease (and strongly implicating tentative immune-based mechanisms) (14-17). Despite these advances, many of these autoantibodies are not yet commercially available, and their predictive utility remains suboptimal, underscoring the need for further validation and refinement of simple and clinically practical biomarkers of SSc-related GI disease.

Although several studies have described the GI manifestations and disease course in SSc, it remains uncertain whether the presence and nature of early GI symptoms can reliably predict future progression (18). A recent clustering analysis using the Medsger GI severity score identified distinct disease trajectories: patients with mild baseline GI involvement and a less severe overall clinical phenotype were less likely to progress (‘stable group’), while others— including early progressive (5%), late progressive (4%), and early severe (4%) groups—were more likely to develop severe GI involvement (9). However, the Medsger severity score does not comprehensively assess GI symptoms; therefore, the range of baseline GI symptoms was not examined. This, unfortunately, limits the ability to differentiate these important patient subgroups in clinical practice. Similarly, another study (19) reported that 4% of patients with diffuse cutaneous SSc developed severe GI disease within 3 years, and 8% within 9 years; however, the initial GI symptoms were not described by the authors, making the early identification of this high-risk subgroup challenging.

Building on this background, we leveraged data from a large, well-characterized cohort of patients with early SSc to assess the presence and severity of baseline GI symptoms and dysfunction. We aimed to determine whether these early features could predict subsequent GI progression and mortality. The main goal was to identify patients whose GI involvement was likely to progress, for clinical care and enrollment in clinical trials.

## Methods

### Study participants

*The* Genetics versus Environment in Scleroderma Outcome Study (GENISOS) cohort, initiated in 1998 (20), is a collaborative effort among the University of Texas Health Science Center at Houston, the University of Texas Medical Branch at Galveston, and the University of Texas Health Science Center at San Antonio. The study enrolled participants aged >18 years who were diagnosed with SSc according to the 1980 American College of Rheumatology (ACR) preliminary classification criteria (21), with subsequent classification using the 2013 American College of Rheumatology/European League Against Rheumatism (EULAR) criteria as these became available (22). Patients were required to have the first non-Raynaud’s symptom attributable to SSc within five years of enrollment. Visits are scheduled approximately every six months during the first three years and annually thereafter. The Institutional Review Boards (IRBs) of all participating institutions approved the study. Written informed consent was obtained from all participants.

### Data collection

A standardized clinical visit form is completed at baseline and follow-up visits. The patient demographic and clinical data collected at the baseline visit included age, sex, race, ethnicity, SSc-specific autoantibody status, disease subtype (diffuse cutaneous or limited cutaneous), and disease duration. Anticentromere antibodies were detected by indirect immunofluorescence using HEp-2 cell substrates (Antibodies, Davis, CA). Anti-topoisomerase I antibody status was determined by passive immunodiffusion against calf thymus extract (Inova Diagnostics, San Diego, CA). Anti–RNA polymerase III antibody status was determined by enzyme-linked immunosorbent assay (MBL, Nagoya, Japan). Modified Rodnan skin score (mRSS) was performed by the rheumatologist examining the patient. The presence or absence of dysphagia (defined as pain or difficulty of swallowing or passing food), gastroesophageal reflux disease (GERD), bloating, constipation, and diarrhea was determined by patient self-report to their physician. A history of peptic ulcer was ascertained by evidence of objective diagnostic testing, malabsorption was defined as diarrhea associated with greater than 10% weight loss or abnormal chemical studies, and intestinal pseudo-obstruction was diagnosed by physician assessment or radiographic evidence. Pulmonary function tests (PFTs) were ordered at the baseline visit, and forced vital capacity percent predicted (FVC%) was used as a surrogate for interstitial lung disease (ILD). GI severity was evaluated using the Medsger GI Severity Score (range 0–4), with higher scores indicating greater disease severity. Scores reflect increasing clinical involvement from no symptoms (0), GERD requiring medication (1), high-dose GERD therapy or antibiotics for bacterial overgrowth (2), pseudo-obstruction or malabsorption (3), to severe dysmotility requiring enteral or parenteral nutrition (4). “Significant GI involvement” was defined as a Medsger GI score ≥ 2 (23). The first instance when the patient reached this score was used to determine the timing of significant GI involvement. Patients’ vital status was determined using medical records and queries obtained from patients’ charts, the National Death Index at the Centers for Disease Control and Prevention, and the Social Security Death Index.

### Statistical analysis

Summary statistics were used to describe baseline patient characteristics. Continuous variables were expressed as mean ± SD, and categorical variables were expressed as % (n). To compare patients who progressed with those who did not, individuals with significant GI disease at baseline and those with only a baseline visit and no follow-up were excluded. A Cox regression model was used to evaluate the association between baseline gastrointestinal manifestations (GERD, dysphagia, bloating, peptic ulcer, constipation, diarrhea) and significant GI severity, defined by a Medsger GI score of 2 or greater. Malabsorption and pseudo-obstruction were excluded from this analysis, as these symptoms indicated that patients already had significant GI severity at baseline. Patients who had these symptoms at baseline were also excluded. For the analysis, time zero was defined as the baseline visit, and participants were censored at their last follow-up visit. Each baseline GI manifestation was included in a univariable model, and then a multivariable model was constructed using the significant predictors from the univariate analysis, along with known confounders identified in the published literature. A separate Cox regression model was also used to evaluate the association between all-cause mortality and baseline GI manifestations, including all manifestations. In mortality analyses, follow-up began at the baseline visit and ended at death or the date of death ascertainment, whichever occurred first. The vital status of each patient was obtained from medical records and patient charts, the National Death Index at the Centers for Disease Control and Prevention, and the Social Security Death Index. In addition, similar models were constructed using the total number of GI manifestations at baseline as the independent variable. Malabsorption and pseudo-obstruction were again excluded from the total symptom count in the analysis involving significant GI severity as the outcome. Sensitivity analyses were also performed, excluding peptic ulcer, as objective diagnostic testing was not available for all participants.

Two-sided p-values less than 0.05 were considered statistically significant. All statistical analyses were performed using STATA/BE 18.0 (StataCorp LP, College Station, TX, USA) or R (version 4.3.2).

## Results

### Characteristics of the patients

A total of 450 patients were included in the study (Table 1). The cohort was predominantly female (84%), with a mean age of 47.7 ± 13.3 years and a mean disease duration of 2.5 ± 1.5 years. Diffuse cutaneous involvement was present in 62% (277/450) of patients. Anti-centromere antibodies were detected in 13% (59/450) of patients, anti–RNA polymerase III antibodies in 21% (94/450), anti-topoisomerase-I antibodies in 18% (82/450), and anti-U3RNP antibodies in 6% (25/450). The mean follow-up time was 5.3 ± 4.8 years with a total of 3041 visits.

**Table 1.** Baseline characteristics of the patients with early systemic sclerosis^a^.

| Variable | Patients<br>n = 450 |
| --- | --- |
| Age, mean ± SD | 47.7 ± 13.3 |
| Female sex, % (n) | 84% (378) |
| Race, % (n) |  |
| White | 77% (345) |
| Black | 19% (85) |
| Asian | 4% (17) |
| Other | 0.7% (3) |
| Hispanic ethnicity, % (n) | 26.7% (120) |
| Disease duration (years), mean ± SD | 2.5 ± 1.5 |
| Diffuse cutaneous involvement, % (n) | 62% (277) |
| Anti-centromere antibody positivity, % (n) | 13% (59) |
| Anti-RNA polymerase III positivity, % (n) | 21% (94) |
| Anti-topoisomerase I positivity, % (n) | 18% (82) |
| Anti-U3RNP positivity, % (n) | 6% (25) |
| GERD, % (n) | 76% (344) |
| Dysphagia, % (n) | 41% (184) |
| Peptic ulcer, % (n) | 5% (21) |
| Bloating, % (n) | 12% (56) |
| Constipation, % (n) | 19% (86) |
| Diarrhea, % (n) | 19% (84) |
| Malabsorption, % (n) | 4% (16) |
| Intestinal pseudo-obstruction, % (n) | 1% (5) |
| Significant GI involvement<sup>b</sup>, % (n) | 4% (19) |
| mRSS, mean ± SD | 16 ± 12 |
| FVC% predicted, mean ± SD | 79.9 ± 19.3 |

Definition of GI symptoms: (a) dysphagia, defined as pain or difficulty in swallowing or passing food; (b) peptic ulcer, confirmed by objective diagnostic testing; (c) malabsorption, defined as diarrhea associated with greater than 10% weight loss or abnormal chemical studies; (d) intestinal pseudo-obstruction, diagnosed by physician assessment or radiographic evidence
<sup>a</sup>Values are the % (n) unless indicated otherwise. FVC: forced vital capacity; GERD: gastroesophageal reflux disease; GI: gastrointestinal; IQR: interquartile range; mRSS: modified Rodnan skin score
<sup>b</sup>Significant GI involvement is defined as a Medsger GI score ≥ 2.

#### At baseline, patients exhibited a range of GI manifestations

GERD was present in 76% patients (344/450), dysphagia in 41% (184/450), constipation in 19% (86/450), diarrhea in 19% (84/450), bloating in 12% (56/450), peptic ulcer in 5% (21/450), malabsorption in 4% (16/450), and pseudo-obstruction in 1% (5/450). Overall, 36.2% patients (163/450) had one manifestation, 27.1% (122/450) had two manifestations, 14.7% (66/450) had three manifestations, 6.9% (31/450) had four manifestations, 2.4% (11/450) had five manifestations, and 0.4% (2/450) had six manifestations. Nobody had seven or eight manifestations at baseline. Twelve percent of patients (55/450) had no GI manifestations at baseline. Among these 55 patients, the maximum number of GI manifestations recorded during follow-up was as follows: 33% patients (18/55) remained free of symptoms (follow-up time: 2.5 ± 2.8 years), while 31% (17/55) developed one manifestation (follow-up time: 7.4 ± 4.9 years), 16% (9/55) developed two (follow-up time: 10.1 ± 5.7 years), 15% (8/55) developed three (follow-up time: 10.5 ± 5.1 years), 4% (2/55) developed four (follow-up time: 12.0 ± 11.3 years), and 2% (1/55) developed five manifestations (follow-up time: 9 years). The mean time to progression to significant GI disease was 4.46 ± 2.65 years.

### GI symptom predictors of significant GI involvement

A total of 13% (58/450) of patients had significant GI involvement, 19 of whom had significant involvement at baseline, while the remainder developed it over the course of follow-up (Figure 1). Among the patients with significant GI disease, 62% (36/58) had a Medsger GI score of 2, 34% (20/58) had a score of 3, and 3% (2/58) had a score of 4 at the time of their first significant GI involvement. We then examined the association between baseline GI manifestations and the risk of developing significant GI severity. Patients who presented with GERD (HR: 4.16, CI: 1.28 – 13.50, *p* = 0.018) or diarrhea (HR: 2.45, CI: 1.24 – 4.85, *p* = 0.010) had a higher risk of progressing to significant GI involvement compared to those who did not (Table 2). The other baseline GI manifestations (i.e., dysphagia, bloating, peptic ulcer, constipation) did not significantly predict progression to significant GI involvement. In a multivariable Cox regression model adjusted for age at enrollment, sex, and disease duration, GERD (HR: 4.15, CI: 1.26 – 13.68, *p* = 0.019) and diarrhea (HR: 2.25, CI: 1.13 – 4.49, *p* = 0.021) remained significantly associated with progression to significant GI involvement. In an analysis using the number of baseline GI manifestations, each additional manifestation was associated with a 48% higher hazard of significant GI severity (HR: 1.48, 95% CI: 1.12 – 1.95, p = 0.005) (Figure 2). This association remained statistically significant (HR: 1.53, 95% CI: 1.16 – 2.02, p = 0.003), even after the model was adjusted for age, sex, and disease duration. For sensitivity analysis, peptic ulcer was removed from the symptom count, and the number of baseline GI manifestations remained significantly associated with significant GI severity in univariable (HR: 1.46, 95% CI: 1.10 – 1.93, p = 0.008) and multivariable analyses (HR: 1.51, 95% CI: 1.14 – 2.00, p = 0.004) (Supplemental Table S2).

**Table 2.**
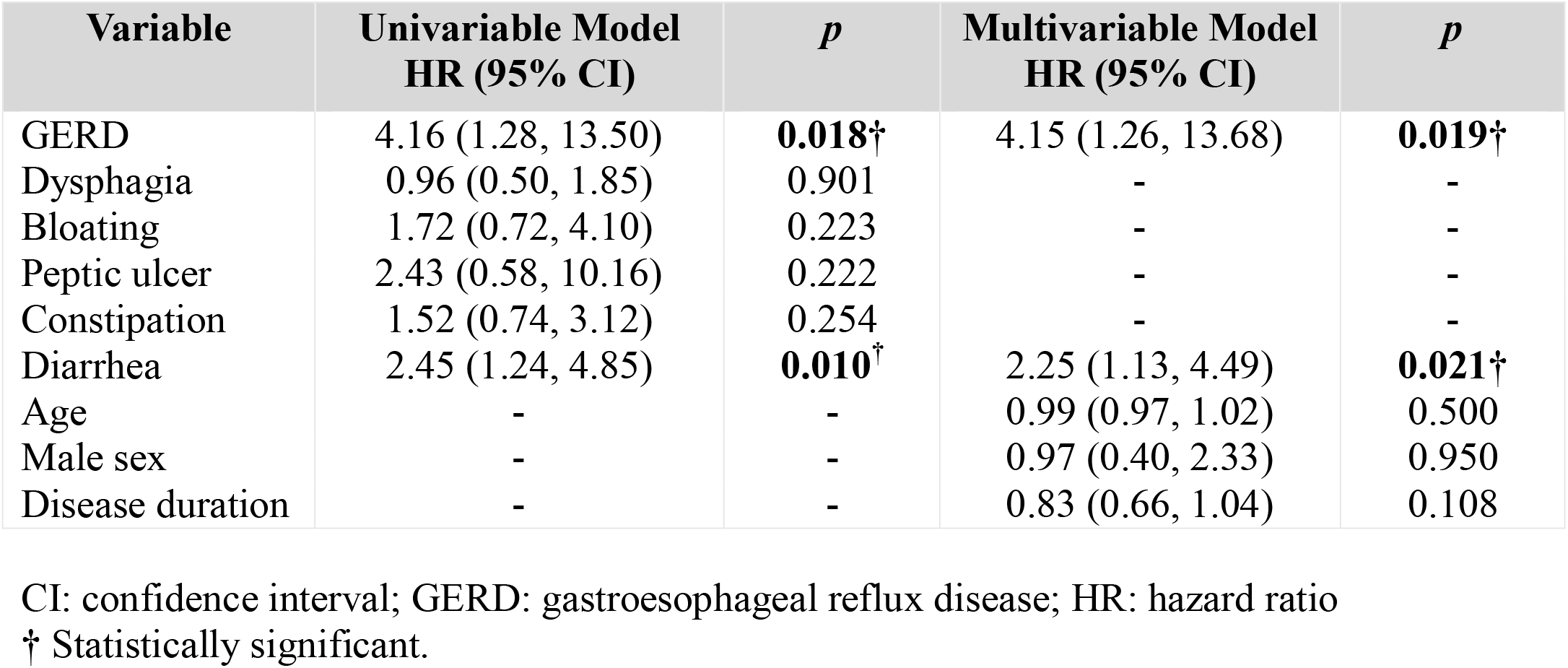
Cox regression models evaluating the association between baseline GI manifestations and time to significant GI disease.

| Variable | Univariable Model<br>HR (95% CI) | *p* | Multivariable Model<br>HR (95% CI) | *p* |
| --- | --- | --- | --- | --- |
| GERD | 4.16 (1.28, 13.50) | **0.018**† | 4.15 (1.26, 13.68) | **0.019**† |
| Dysphagia | 0.96 (0.50, 1.85) | 0.901 | - | - |
| Bloating | 1.72 (0.72, 4.10) | 0.223 | - | - |
| Peptic ulcer | 2.43 (0.58, 10.16) | 0.222 | - | - |
| Constipation | 1.52 (0.74, 3.12) | 0.254 | - | - |
| Diarrhea | 2.45 (1.24, 4.85) | **0.010**† | 2.25 (1.13, 4.49) | **0.021**† |
| Age | - | - | 0.99 (0.97, 1.02) | 0.500 |
| Male sex | - | - | 0.97 (0.40, 2.33) | 0.950 |
| Disease duration | - | - | 0.83 (0.66, 1.04) | 0.108 |

CI: confidence interval; GERD: gastroesophageal reflux disease; HR: hazard ratio
† Statistically significant.

**Figure 1.**
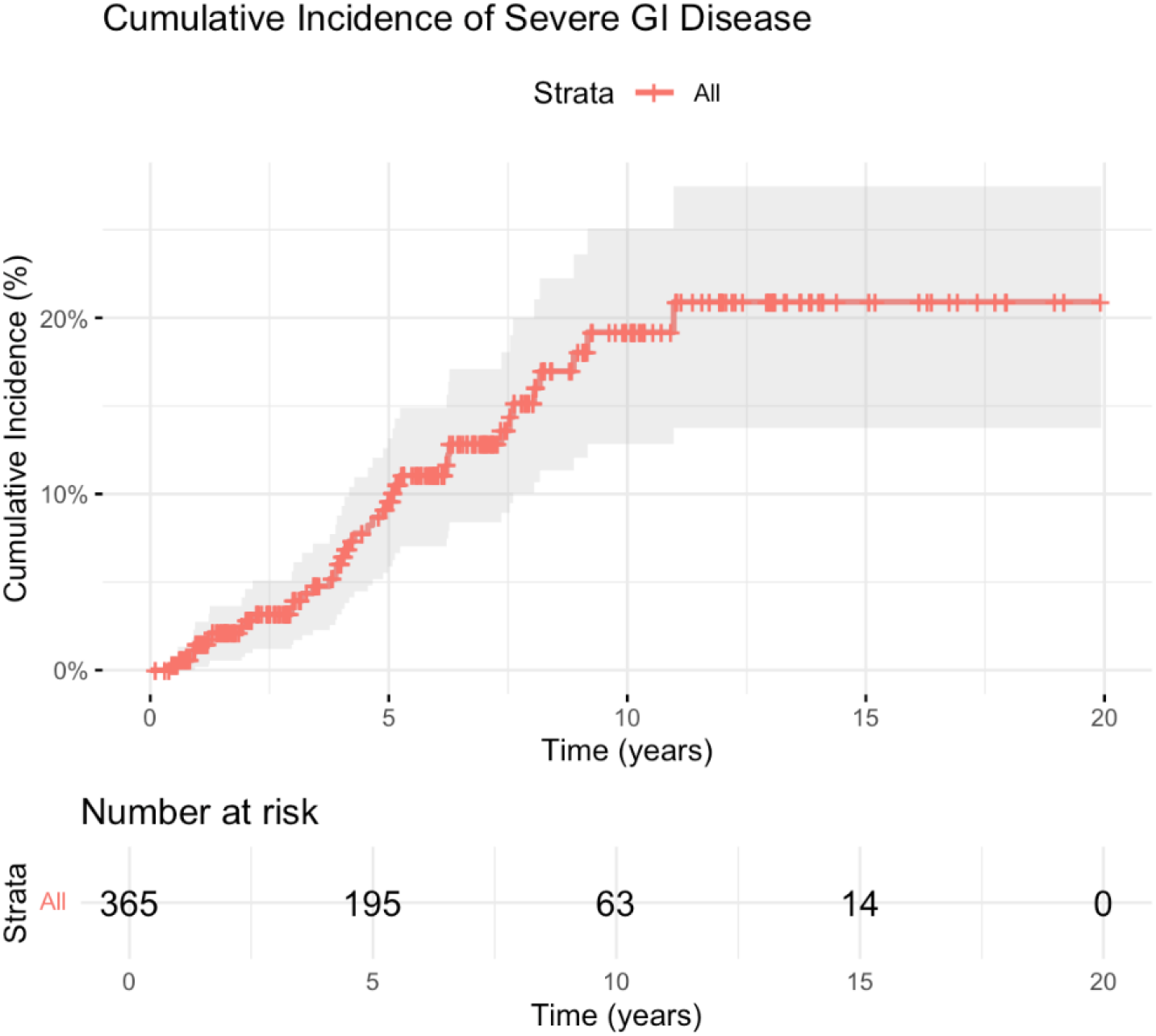
Cumulative incidence curve showing cumulative incidence of significant GI disease (Medsger GI score ≥2) progression. Cumulative incidence curve showing estimated cumulative incidence of progression to significant GI disease (Medsger GI score ≥2) among patients without significant GI involvement at baseline. The red step function represents the estimated cumulative incidence over time, with shaded areas indicating 95% confidence intervals. Marks on the curve denote censoring events. The number-at-risk table below illustrates the number of participants who remained under observation at each time point. Overall, approximately 20% of patients progressed to significant GI disease during follow-up, with increasing risk observed over the first 10 years.

**Figure 2.**
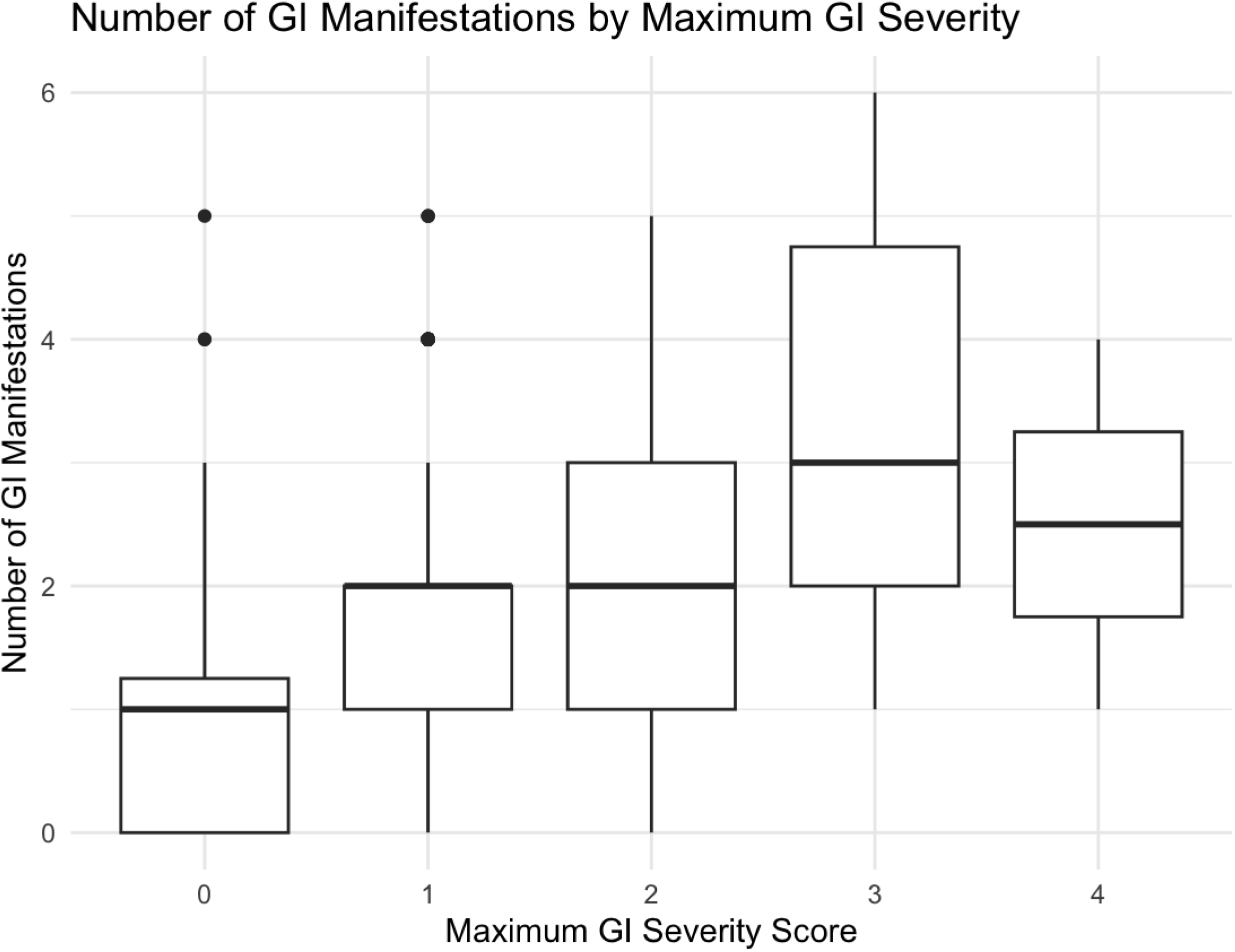
Number of GI manifestations at baseline by maximum GI severity score ever recorded. Boxplot showing the number of baseline GI manifestations—including dysphagia, gastroesophageal reflux disease (GERD), bloating, constipation, diarrhea, peptic ulcer, malabsorption, and pseudo-obstruction—across categories of maximum GI severity recorded during follow-up. Each box represents the median, interquartile range, and distribution of symptom counts within each severity level, with whiskers indicating variability and dots marking outliers. Higher GI severity scores are associated with a greater number of GI manifestations, demonstrating increasing symptom burden with more advanced GI involvement.

### Predictors of mortality

We further examined the association between individual baseline GI manifestations and all-cause mortality. Patients who presented with peptic ulcer (HR: 2.18, CI: 1.24 – 3.83, *p* = 0.007), diarrhea (HR: 1.51, CI: 1.07 – 2.13, *p* = 0.018), or malabsorption (HR: 2.53, CI: 1.37 – 4.66, *p* = 0.003) had a higher risk of mortality compared to those who did not (Table 3). The other GI manifestations (i.e., GERD, dysphagia, bloating, constipation, pseudo-obstruction) did not significantly predict mortality. In a multivariable model adjusted for age, sex, and disease duration, diarrhea (HR: 1.48, CI: 1.01 – 2.18, *p* = 0.045) and peptic ulcer (HR: 2.21, CI: 1.25 – 3.91, *p* = 0.007) remained significantly associated with higher mortality, whereas the association with malabsorption lost statistical significance. Finally, using a Cox proportional hazards model to evaluate the number of baseline GI manifestations (i.e., baseline GI symptom burden), we found that <u>each</u> additional manifestation was associated with a 26% higher hazard of all-cause mortality (HR: 1.26, 95% CI: 1.11 – 1.42, p < 0.001). This association remained significant (HR: 1.29, 95% CI: 1.14 – 1.45, p < 0.001) after adjusting the model for age, sex, and disease duration (Table 4). For sensitivity analysis, peptic ulcer was removed from the symptom count, and the number of baseline GI manifestations remained significantly associated with mortality in univariable (HR: 1.23, 95% CI: 1.09 – 1.40, p < 0.001) and multivariable analyses (HR: 1.26, 95% CI: 1.12 – 1.43, p < 0.001) (Supplemental Table S2).

**Table 3.**
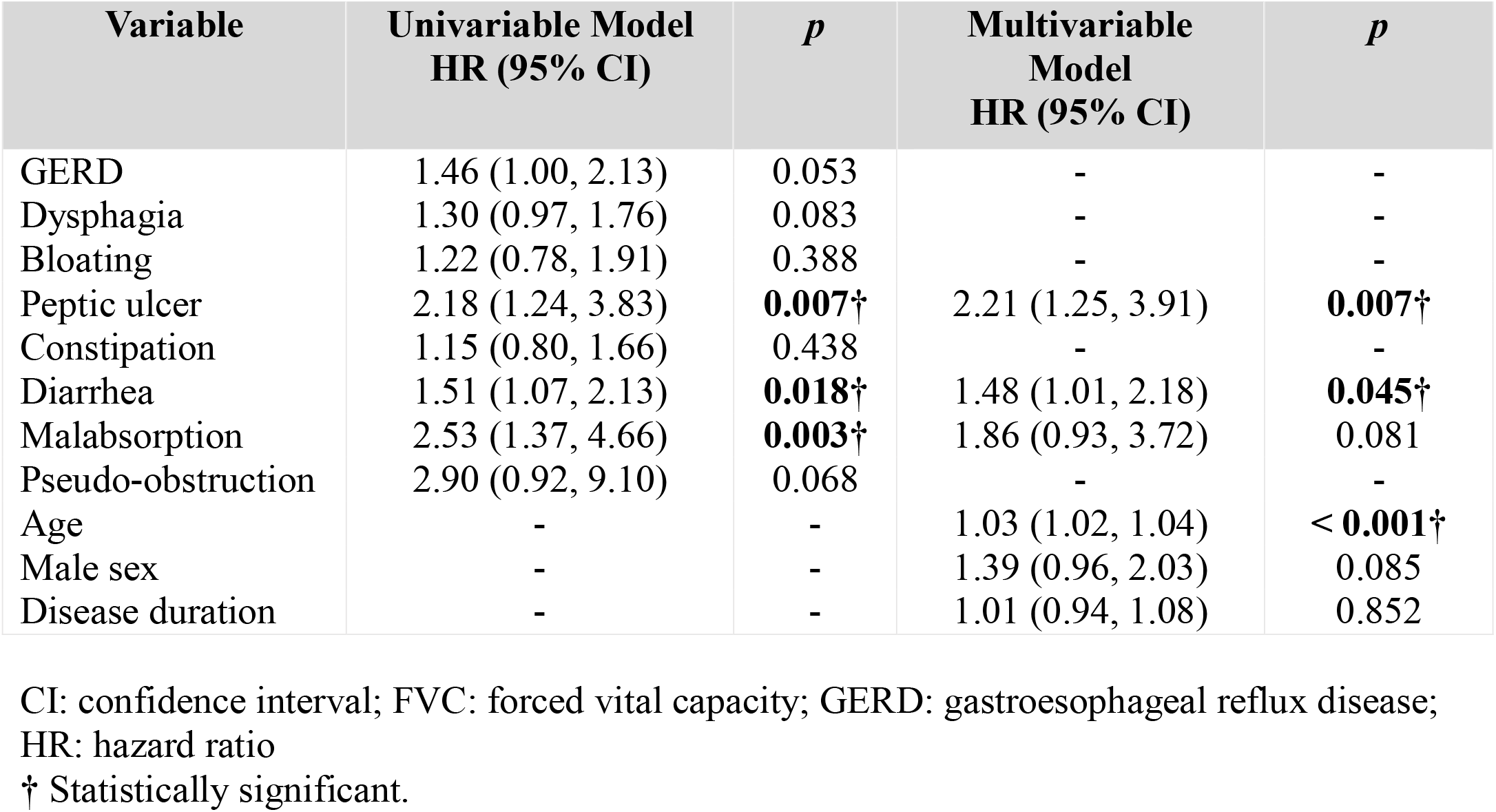
Cox regression models evaluating the association between baseline GI manifestations and time to death.

| Variable | Univariable Model<br>HR (95% CI) | *p* | Multivariable Model<br>HR (95% CI) | *p* |
| --- | --- | --- | --- | --- |
| GERD | 1.46 (1.00, 2.13) | 0.053 | - | - |
| Dysphagia | 1.30 (0.97, 1.76) | 0.083 | - | - |
| Bloating | 1.22 (0.78, 1.91) | 0.388 | - | - |
| Peptic ulcer | 2.18 (1.24, 3.83) | **0.007**† | 2.21 (1.25, 3.91) | **0.007**† |
| Constipation | 1.15 (0.80, 1.66) | 0.438 | - | - |
| Diarrhea | 1.51 (1.07, 2.13) | **0.018**† | 1.48 (1.01, 2.18) | **0.045**† |
| Malabsorption | 2.53 (1.37, 4.66) | **0.003**† | 1.86 (0.93, 3.72) | 0.081 |
| Pseudo-obstruction | 2.90 (0.92, 9.10) | 0.068 | - | - |
| Age | - | - | 1.03 (1.02, 1.04) | **< 0.001**† |
| Male sex | - | - | 1.39 (0.96, 2.03) | 0.085 |
| Disease duration | - | - | 1.01 (0.94, 1.08) | 0.852 |

CI: confidence interval; FVC: forced vital capacity; GERD: gastroesophageal reflux disease; HR: hazard ratio
† Statistically significant.

**Table 4.** Multivariable Cox Regression model for symptom count predicting significant GI disease and time to death.

| Variable | Time to Significant GI Disease (HR, 95% CI) | p | Time to Death (HR, 95% CI) | p |
| --- | --- | --- | --- | --- |
| Symptom count | 1.53 (1.16–2.02) | **0.003**† | 1.29 (1.14–1.45) | **<0.001**† |
| Male sex | 1.00 (0.42–2.43) | 0.993 | 1.54 (1.06–2.23) | **0.022**† |
| Age | 0.99 (0.97–1.02) | 0.514 | 1.03 (1.02–1.04) | **<0.001**† |
| Disease duration | 0.83 (0.66–1.05) | 0.118 | 1.01 (0.93–1.08) | 0.892 |

CI: confidence interval
† Statistically significant.

## Discussion

GI disease in SSc presents with remarkable variability within individual patients, a feature that complicates clinical management but also offers an important opportunity to improve risk stratification and personalized care. In this study, we demonstrated that patients with SSc have a range of GI manifestations of varying frequency and, for the first time in a large prospective cohort of patients with early SSc, used these patterns of clinical presentation to predict later GI progression and mortality. These findings highlight the potential to use symptom profiles as predictors to inform routine clinical care and enrich clinical trial enrollment, particularly given the limited availability of validated commercial biomarkers for SSc-related GI disease.

GERD was the only baseline symptom present in the majority of patients, occurring in 76% at baseline, followed by the next most common symptom, dysphagia, which was present at baseline in 41% of patients. This finding is consistent with previous literature reporting a prevalence of esophageal involvement greater than 90% in patients with SSc (6). One of the striking findings of our study was that patients who presented with GERD had more than a fourfold increased risk of progressing to significant GI involvement compared to those without GERD, even after adjusting for age, sex, and disease duration. In other words, not having GERD at baseline was associated with a 76% lower risk of developing significant GI disease over time. This finding suggests that the presence of GERD at baseline may serve as an important clinical indicator of future GI involvement in SSc, and that patients without GERD are, on average, less likely to develop significant GI disease over the course of their illness.

We also demonstrated that diarrhea at baseline was associated with a 2.5-fold increased risk of developing significant GI disease in patients with SSc. In our cohort, the baseline prevalence of diarrhea was 19%, the same as that of constipation. This suggests that diarrhea at baseline, compared to constipation, may confer a relatively higher risk of progression to severe GI involvement. Given the wide range of diarrhea severity (24), clinicians should recognize that even diarrhea without malabsorption can signal a higher likelihood of future GI progression and complications in SSc.

In our analysis using the number of baseline GI manifestations, we found that each additional GI manifestation was associated with a 48% higher risk of developing significant GI severity. This finding indicates that a greater GI burden (i.e., a higher number of manifestations) at baseline portends a worse GI course for patients with SSc. A recent study (25) showed that the number of GI segments involved throughout the disease was associated with distinct GI disease characteristics; however, we are the first to use baseline GI symptom burden (i.e., count) to predict progression and outcomes. These observations suggest that physicians should carefully assess (and document) both the presence and extent of baseline GI manifestations in patients presenting with SSc and consider using this information to help stratify risk.

In our mortality analyses, malabsorption was associated with an approximately 2.5-fold increased risk of death, peptic ulcer disease with a 2.2-fold increased risk, and diarrhea with a more modest 1.5-fold increase in risk. These findings underscore the importance of GI involvement in SSc outcomes. Notably, a recent meta-analysis (26) reported a higher prevalence of *Helicobacter pylori* infection in patients with SSc compared to healthy controls, highlighting a potential infectious contributor to the development of peptic ulcers. Moreover, vascular and autonomic dysfunction, which are hallmarks of SSc pathophysiology (27), may further predispose patients to peptic ulcers, as autonomic dysfunction has been proposed as a necessary condition for their development (28). Despite its clinical relevance, autonomic dysfunction remains an important yet often overlooked aspect of SSc. Diarrhea and malabsorption also carry profound implications, as they can lead to malnutrition, which is a known predictor of increased mortality in SSc (29). Additionally, we observed that patients with a higher baseline GI symptom burden were more likely to experience adverse outcomes. Taken together, these findings suggest that patients presenting with peptic ulcer disease, diarrhea, malabsorption, or multiple GI manifestations at baseline should be considered at ‘high-risk’ and warrant closer clinical monitoring and frequency of follow-up.

This study benefits from several key strengths, including a well-characterized cohort of patients with early SSc, long-term follow-up, and comprehensive documentation of GI symptoms. The inclusion of a diverse patient population further enhances the generalizability of the findings. However, several limitations should be acknowledged. The use of all-cause mortality as the primary outcome, while clinically meaningful, does not explicitly capture GI-related deaths; nonetheless, this approach successfully identified a subset of patients with a high-risk clinical phenotype. Additionally, the total gastrointestinal symptom count weighted all manifestations equally, potentially oversimplifying heterogeneity in symptom severity and prognostic relevance. Not all patients underwent objective GI evaluations, which may have led to underdiagnosis of asymptomatic ulcers or other GI complications. Finally, the lack of objective assessments of GI motility limited our ability to characterize the extent of GI involvement fully.

Our findings demonstrate that baseline GI manifestations, including peptic ulcer disease, diarrhea, and malabsorption, are associated with severe GI outcomes and increased mortality in patients with SSc. These results have important implications for risk stratification in both clinical practice and for enrollment in clinical trials, particularly in light of the limited availability of commercial biomarkers to guide prognostication for GI disease in this population.

## Acknowledgements

Author [M.H.] is supported by the NIHR Manchester Biomedical Research Centre (NIHR203308).

## Data availability statement

The data underlying this article will be shared on reasonable request to the corresponding author.

## References

1. Denton CP, Khanna D. Systemic sclerosis. Lancet. 2017;390(10103):1685–99.

2. Li JX. Secular Trends in Systemic Sclerosis Mortality in the United States from 1981 to 2020. Int J Environ Res Public Health. 2022;19(22).

3. Distler O, Assassi S, Cottin V, Cutolo M, Danoff SK, Denton CP, et al. Predictors of progression in systemic sclerosis patients with interstitial lung disease. Eur Respir J. 2020;55(5).

4. van Leeuwen NM, Maurits M, Liem S, Ciaffi J, Ajmone Marsan N, Ninaber M, et al. New risk model is able to identify patients with a low risk of progression in systemic sclerosis. RMD Open. 2021;7(2).

5. Herrick AL, Peytrignet S, Lunt M, Pan X, Hesselstrand R, Mouthon L, et al. Patterns and predictors of skin score change in early diffuse systemic sclerosis from the European Scleroderma Observational Study. Ann Rheum Dis. 2018;77(4):563–70.

6. McMahan ZH, Kulkarni S, Chen J, Chen JZ, Xavier RJ, Pasricha PJ, et al. Systemic sclerosis gastrointestinal dysmotility: risk factors, pathophysiology, diagnosis and management. Nat Rev Rheumatol. 2023;19(3):166–81.

7. Dein E, Kuo PL, Hong YS, Hummers LK, Mecoli CA, McMahan ZH. Evaluation of risk factors for pseudo-obstruction in systemic sclerosis. Semin Arthritis Rheum. 2019;49(3):405–10.

8. McMahan ZH, Paik JJ, Wigley FM, Hummers LK. Determining the Risk Factors and Clinical Features Associated With Severe Gastrointestinal Dysmotility in Systemic Sclerosis. Arthritis Care Res (Hoboken). 2018;70(9):1385–92.

9. Perin J, Hughes M, Mecoli CA, Paik JJ, Gelber AC, Wigley FM, et al. Distinct clinical trajectories of gastrointestinal progression among patients with systemic sclerosis. Rheumatology (Oxford). 2025;64(5):2766–74.

10. Tucker AE, Perin J, Volkmann ER, Abdi T, Shah AA, Pandolfino J, et al. Associations Between Patterns of Esophageal Dysmotility and Extra-Intestinal Features in Patients With Systemic Sclerosis. Arthritis Care Res (Hoboken). 2023;75(8):1715–24.

11. Salas AD, Yanek LR, Hummers LK, Shah AA, McMahan ZH. Abnormal Esophageal Scintigraphy Associates With a Distinct Clinical Phenotype in Patients With Systemic Sclerosis. ACR Open Rheumatol. 2025;7(1):e11796.

12. Cheah JX, Perin J, Hughes M, Mecoli CA, Paik JJ, Hummers LK, et al. Demographics and clinical features associated with abnormal small bowel motility in systemic sclerosis. Rheumatology (Oxford). 2025;64(5):2775–82.

13. Cheah JX, Perin J, Volkmann ER, Hummers LK, Pasricha PJ, Wigley FM, et al. Slow Colonic Transit in Systemic Sclerosis: An Objective Assessment of Risk Factors and Clinical Phenotype. Arthritis Care Res (Hoboken). 2023;75(2):289–98.

14. Ayla AY, Kalavar NR, Pimentel M, Morales W, Hummers LK, Shah AA, et al. Antimuscarinic 3 antibodies associate with a severe clinical phenotype in patients with systemic sclerosis. Rheumatology (Oxford). 2025;64(10):5230–7.

15. McMahan ZH, Puttapaka SN, Casciola-Rosen L, Kaniecki T, Gutierrez-Alamillo L, Ming SH, et al. Antimitochondrial antibodies in systemic sclerosis target enteric neurons and are associated with GI dysmotility. Ann Rheum Dis. 2025;84(10):1721–32.

16. Herrán M, Adler BL, Perin J, Morales W, Pimentel M, McMahan ZH. Antivinculin Antibodies in Systemic Sclerosis: Associations With Slow Gastric Transit and Extra-intestinal Clinical Phenotype. Arthritis Care Res (Hoboken). 2023;75(10):2166–73.

17. McMahan ZH, Kulkarni S, Andrade F, Perin J, Zhang C, Hooper JE, et al. Anti-Gephyrin Antibodies: A Novel Specificity in Patients With Systemic Sclerosis and Lower Bowel Dysfunction. Arthritis Rheumatol. 2024;76(1):92–9.

18. Hughes M, Sylvestre Y, Manning J, Wragg E, Mandzuk M, Samaranayaka M, et al. Longitudinal trajectories of the Scleroderma Health Assessment Questionnaire (SHAQ) visual analogue scales: a retrospective cohort study. Rheumatology (Oxford). 2025;64(5):2821–7.

19. Steen VD, Medsger TA, Jr. Severe organ involvement in systemic sclerosis with diffuse scleroderma. Arthritis Rheum. 2000;43(11):2437–44.

20. Reveille JD, Fischbach M, McNearney T, Friedman AW, Aguilar MB, Lisse J, et al. Systemic sclerosis in 3 US ethnic groups: a comparison of clinical, sociodemographic, serologic, and immunogenetic determinants. Semin Arthritis Rheum. 2001;30(5):332–46.

21. Preliminary criteria for the classification of systemic sclerosis (scleroderma). Subcommittee for scleroderma criteria of the American Rheumatism Association Diagnostic and Therapeutic Criteria Committee. Arthritis Rheum. 1980;23(5):581–90.

22. van den Hoogen F, Khanna D, Fransen J, Johnson SR, Baron M, Tyndall A, et al. 2013 classification criteria for systemic sclerosis: an American college of rheumatology/European league against rheumatism collaborative initiative. Ann Rheum Dis. 2013;72(11):1747–55.

23. McMahan ZH. Gastrointestinal involvement in systemic sclerosis: an update. Curr Opin Rheumatol. 2019;31(6):561–8.

24. Sakkas LI, Simopoulou T, Daoussis D, Liossis SN, Potamianos S. Intestinal Involvement in Systemic Sclerosis: A Clinical Review. Dig Dis Sci. 2018;63(4):834–44.

25. Mollière C, Guédon AF, Kapel N, de Vassoigne F, Graiess F, Senet P, et al. Gastrointestinal disorders in systemic sclerosis: cluster analysis and prognosis from a French prospective cohort. Clin Exp Rheumatol. 2024;42(8):1656–64.

26. Yong WC, Upala S, Sanguankeo A. Helicobacter pylori infection in systemic sclerosis: a systematic review and meta-analysis of observational studies. Clin Exp Rheumatol. 2018;36 Suppl 113(4):168–74.

27. Alvarez-Hernandez MP, Adler B, Perin J, Hughes M, McMahan ZH. Evaluating the associations between dysautonomia, gastrointestinal transit, and clinical phenotype in patients with systemic sclerosis. Arthritis Care Res (Hoboken). 2024.

28. Nomura M, Yukinaka M, Miyajima H, Nada T, Kondo Y, Okahisa T, et al. Is autonomic dysfunction a necessary condition for chronic peptic ulcer formation? Aliment Pharmacol Ther. 2000;14 Suppl 1:82–6.

29. Assassi S, Del Junco D, Sutter K, McNearney TA, Reveille JD, Karnavas A, et al. Clinical and genetic factors predictive of mortality in early systemic sclerosis. Arthritis Rheum. 2009;61(10):1403–11.

