## Supplemental Table 1 for "Early gastrointestinal manifestations predict disease progression and mortality in patients with systemic sclerosis"

**Supplementary Table S1. Baseline characteristics of gastrointestinal progressors vs. non-progressors^a^**

| **Variable** | **Patients**  **Non-progressors (n = 326)** | **Patients**  **Progressors (n = 39)** | ***p*** |
| --- | --- | --- | --- |
| Age, mean ± SD | 48.7 ± 13.6 | 47.0 ± 11.7 | 0.400 |
| Female sex, % (n) | 83% (271) | 85% (33) | 0.994 |
| Race, % (n)  White  Black  Asian  Other | 78% (254)  17% (55)  5% (15)  0.6% (2) | 72% (28)  28% (11)  0  0 | 0.220 |
| Hispanic ethnicity, % (n) | 26% (85) | 13% (5) | 0.078 |
| Disease duration (years), mean ± SD | 2.6 ± 2.2 | 2.2 ± 1.3 | 0.109 |
| Follow-up time (years), mean ± SD | 6.0 (2.5 – 10.0) | 8.0 (5.5-12.0) | **0.009**† |
| Diffuse cutaneous involvement, % (n) | 59% (193) | 67% (26) | 0.468 |
| Anti-centromere antibody positivity | 15% (50/323) | 8% (3/39) | 0.237 |
| Anti-RNA polymerase III positivity | 22% (71/319) | 23% (9/39) | 1.000 |
| Anti-topoisomerase I positivity | 19% (61/323) | 10% (4/39) | 0.268 |
| Anti-U3RNP positivity | 6% (15/237) | 12% (4/32) | 0.258 |
| GERD, % (n) | 72% (235) | 92% (36) | **0.006**† |
| Dysphagia, % (n) | 37% (120) | 36% (14) | 1.000 |
| Peptic ulcer, % (n) | 4% (12) | 5% (2) | 0.652 |
| Bloating, % (n) | 10% (32) | 15% (6) | 0.424 |
| Constipation, % (n) | 17% (56) | 26% (10) | 0.281 |
| Diarrhea, % (n) | 15% (48) | 31% (12) | **0.020**† |
| mRSS, mean ± SD | 15.8 ± 11.7 | 18.8 ± 12.0 | 0.142 |
| FVC% predicted, mean ± SD | 82.0 ± 18.6 | 78.9 ± 20.4 | 0.400 |

^a^ Progression was defined as an increase in gastrointestinal (GI) disease severity, indicated by a Medsger GI score of ≥2 at any point following the baseline visit, reflecting the development of clinically significant GI involvement.

† Statistically significant.
