## Supplemental Table 2 for "Early gastrointestinal manifestations predict disease progression and mortality in patients with systemic sclerosis"

**Supplementary Table S2.** **Multivariable Cox Regression model for symptom count predicting significant GI disease and time to death, excluding peptic ulcer from symptom count**

| **Variable** | **Time to Significant GI Disease (HR, 95% CI)** | **p** | **Time to Death (HR, 95% CI)** | **p** |
| --- | --- | --- | --- | --- |
| Symptom count | 1.51 (1.14–2.00) | **0.004**† | 1.26 (1.12–1.43) | **<0.001**† |
| Male sex | 1.00 (0.41–2.40) | 0.991 | 1.53 (1.06–2.21) | **0.024**† |
| Age | 0.99 (0.97–1.02) | 0.521 | 1.03 (1.02–1.04) | **<0.001**† |
| Disease duration | 0.84 (0.67–1.05) | 0.121 | 1.00 (0.93–1.08) | 0.918 |
